# Cost-effectiveness of the Strategies for Surveillance of Antimicrobial-resistant Gonorrhea in the US: a Modelling Study

**DOI:** 10.1101/2024.07.29.24311166

**Authors:** Sofya Prakhova

## Abstract

**Background:** The Gonococcal Isolate Surveillance Project (GISP) is a sentinel surveillance system to monitor the spread of antimicrobial-resistant (AMR) gonorrhea. Under GISP surveillance strategy, urethral isolates are utilized for monitoring the spread of the resistance and the obtained estimates are used for informing the gonorrhea treatment guidelines. In 2017, the enhanced Gonococcal Isolate Surveillance Project (eGISP) was established which also includes the non-urethral isolates. Using eGISP estimates for informing the gonorrhea treatment guidelines is an alternative surveillance strategy that can be used.

**Methods:** We utilized our previously developed continuous-time agent-based model of gonorrhea transmission among the US men who have sex with men (MSM) population and calculated the total number of discounted quality-adjusted life years (QALYs) and total discounted costs over 25 years under GISP and eGISP surveillance strategy. We also evaluated cost-effectiveness of both surveillance strategies.

**Results:** Under GISP surveillance strategy, $2.9M (95% uncertainty interval: $23,131, $9.4M) were saved and 31.3 (0, 134.9) QALYs were gained in the simulated cohort of 10,000 US MSM over 25 years compared to no surveillance. Performing eGISP surveillance strategy instead would result in additional $57,449 (−$100,914, $221,663) saved and 0.59 (−0.79, 2.5) QALYs gained.

**Conclusion:** The current GISP surveillance strategy significantly reduces the costs and increases the health benefits compared to no surveillance. However, switching from the current strategy to eGISP strategy is cost saving and should be considered in order to improve the population health and reduce the financial burden of gonorrhea.

## Introduction

*N. gonorrhoeae* has developed resistance over the years to most classes of antibiotic used for its treatment (penicillins, sulphonamides, tetracyclines, quinolones and macrolide) and continues to develop resistance to the so-called last-line cephalosporins [1], making it an urgent global public health threat. Untreated gonorrhea can have a number of health consequences such as infertility, disseminated gonococcal infection (DGI), ectopic pregnancy, increased ability to receive and transmit HIV etc. [2]. In addition to negative impact on health, some of these complications involve high financial costs for both individuals and health systems.

The World Health Organization (WHO) has defined a number of measures that can fight antimicrobial-resistant (AMR) gonorrhea, including establishing effective drug regulations, strengthening surveillance systems for antimicrobial resistance, support of research to find low-cost tests to identify *N. gonorrhoeae* etc. Promising future options are point-of-care tests and gonococcal vaccine [3] [4]. However, until they are widespread, the goal is to allocate the existing resources optimally to the existing options.

In the US, the gonorrhea treatment guidelines are based on the estimates from the Gonococcal Isolate Surveillance Project (GISP) [5], a sentinel surveillance system that utilizes the urethral isolates and estimates the percentage of diagnosed cases resistant to antibiotics currently or previously used for the treatment of gonorrhea. Once that value reaches 5% for the current fist-line antibiotic, a switch to a different drug is meant to be made according to the WHO guidelines [6]. In 2017, the enhanced Gonococcal Isolate Surveillance Project (eGISP) [7] was established, which includes the extragenital isolates in addition to the urethral ones used in GISP.

In our earlier work [8], we evaluated performance of both surveillance systems. It was determined that eGISP system detects the moment of switch to a new first-line drug more accurately than the original system which results in lower number of gonorrhea cases without reduction in the lifespan of the current first-line antibiotic. However, the costs involved remain unknown.

In this paper we evaluate cost-effectiveness of GISP and eGISP surveillance strategies (utilizing GISP or eGISP estimates for informing the gonorrhea treatment guidelines). In order to do this, we utilize our previously developed agent-based model of gonorrhea transmission among the US men who have sex with men (MSM) and estimate the total number of discounted quality-adjusted life years (QALYs) and total discounted costs under both surveillance strategies over a 25-year period. The contribution of different types of gonococcal infection and of the sequalae into the overall disease burden under GISP and eGISP strategy was also investigated.

## Methods

### Simulation model and surveillance strategies

The continuous-time agent-based model of gonorrhea transmission among the US MSM is described in details in [8]. The population of the model is 10,000. The model accounts for susceptible and resistant strains of gonorrhea. The infection can be either symptomatic or asymptomatic. It can be transmitted between rectum and pharynx, rectum and urethra, pharynx and urethra, pharynx and pharynx, and urethra and urethra. A one-site and two-site infection is allowed.

Symptomatic individuals seek the first-line treatment (ceftriaxone) at healthcare facilities, while asymptomatic ones either recover naturally or get detected during the screening and receive the treatment. The treatment fails if individuals are infected with resistant strain of gonorrhea or if the bacteria develops resistant to ceftriaxone during the treatment. In this case, symptomatic individuals are being re-treated with the second-line drug (ertapenem), while asymptomatic ones remain infectious.

The model was calibrated using the Bayesian calibration approach to the local epidemiological data: prevalence of gonorrhea among the US MSM at three anatomical sites, prevalence of gonorrhea resistant to ceftriaxone among the US MSM at three anatomical sites and the annual rate of reported gonorrhea cases per 100,000 US MSM population. The model was developed using a simulation modelling tool AnyLogic (version 8.8.1 University).

A surveillance system for monitoring the spread of AMR gonorrhea (GISP or eGISP) was modelled as a sum of the percentage of diagnosed cases resistant to ceftriaxone and the estimation error due to the fact that only a limited number of isolates is tested for drug susceptibility each year. Once a surveillance system detects that the percentage of cases resistant to ceftriaxone has reached 5%, a switch to a different drug is made according to the WHO guidelines [6].

Under GISP surveillance strategy, urethral isolates from the first 25 men diagnosed with urethral gonorrhea at a number of surveillance sites are utilized for estimating the percentage of cases resistant to ceftriaxone. The obtained estimates are used for informing the gonorrhea treatment guidelines. Under eGISP surveillance strategy, urethral, rectal and pharyngeal isolates are applied for this purpose. This is an alternative surveillance strategy that can be applied.

### Cost-effectiveness analysis

We adapted a healthcare sector perspective for our analysis and followed the Consolidated Health Economic Evaluation Reporting Standards (CHEERS) [9]. We used QALYs as a measure of the disease burden on the population health as it allows to include morbidity and the impact of sequalae due to gonorrhea, and to compare the efficiency of health interventions for different conditions using the same units.

It was assumed that people experience reduced quality of life due to urethritis during symptomatic urethral infection and while experiencing sequalae due to untreated asymptomatic gonorrhea. In case of symptomatic resistant infection, we assumed that individuals experience urethritis for longer as at first, they are treated unsuccessfully with the first-line drug. We considered epididymitis and disseminated gonococcal infection (DGI) as possible sequalae and distinguished between inpatient and outpatient treatment of these sequalae. The probability of developing a sequalae was assumed to be the same for all anatomical sites and independent of the infection duration as there is limited evidence on the relationships between them [10].

The state specific utilities, probabilities and durations as well as the associated uncertainty intervals were obtained from the earlier comprehensive study of gonorrhea burden [11] and are presented in Table 1.

**Table 1.** Utilities, probabilities and durations used in the analysis. Abbreviation: DGI, disseminated gonococcal infection.

| Parameter | Mean value | Uncertainty interval | Source |
| --- | --- | --- | --- |
| **Utilities** |  |  | [11] |
| Urethritis | 0.85 | 0.76 to 0.92 |  |
| DGI (inpatient treatment) | 0.68 | 0.53 to 0.84 |  |
| DGI (outpatient treatment) | 0.60 | 0.41 to 0.79 |  |
| Epididymitis (inpatient treatment) | 0.30 | 0.012 to 0.59 |  |
| Epididymitis (outpatient treatment) | 0.46 | 0.20 to 0.72 |  |
| **Probabilities** |  |  | [11] |
| Probability of inpatient treatment given DGI | 0.29 | 0.17 to 0.43 |  |
| Probability of inpatient treatment given epididymitis | 0.0054 | 0.0028 to 0.0092 |  |
| Probability of DGI given untreated gonorrhea | 0.01 | 0.0075 to 0.013 |  |
| Probability of epididymitis given untreated gonorrhea | 0.042 | 0.0012 to 0.14 |  |
| **Durations (years)** |  |  | [11] |
| Urethritis | 0.019 | 0.01 to 0.028 |  |
| DGI (inpatient treatment) | 0.030 | 0.016 to 0.044 |  |
| DGI (outpatient treatment) | 0.022 | 0.011 to 0.032 |  |
| Epididymitis (inpatient treatment) | 0.0082 | 0.0043 to 0.012 |  |
| Epididymitis (outpatient treatment) | 0.019 | 0.0010 to 0.028 |  |

The total costs were calculated as the sum of the costs due to diagnosis and treatment of gonorrhea and the costs of treatment of the sequelae. We distinguished between diagnosis and treatment of symptomatic versus asymptomatic gonococcal infection as well as susceptible strain of gonorrhea versus ceftriaxone-resistant strain. All the costs are listed in Table 2. The costs were adjusted to 2017 USD using the medical care component of the consumer price index [12]. The details of calculation are provided in the supplement.

**Table 2.** Costs used in the analysis. Abbreviation: DGI, disseminated gonococcal infection.

| Costs (in 2017 USD) | Mean value | Uncertainty interval | Source |
| --- | --- | --- | --- |
| Testing for gonorrhea | 65.4 | 34.2 to 97.6 | [11] |
| Treatment of urethritis | 92.4 | 74.4 to 110.5 | [11] |
| Short clinic visit | 41.5 | 32.9 to 49.9 | [11] |
| Ceftriaxone 250g | 24.5 | 19 to 29.6 | [11] |
| Azithromycin 1g | 30.9 | 24.4 to 37.2 | [11] |
| Ertapenem 1g | 64.6 | 42.5 to 86.6 | [13] |
| Treatment of DGI (inpatient) | 7,694.5 | 6,155.2 to 9,233.9 | [11] |
| Treatment of DGI (outpatient) | 632.5 | 506 to 758.9 | [11] |
| Treatment of epididymitis (inpatient) | 8,302.5 | 6,642 to 9,970.5 | [11] |
| Treatment of epididymitis (outpatient) | 475.2 | 378.4 to 569.7 | [11] |

The simulations were performed for 25 years starting from the year when eGISP was established (2017). This simulation window was chosen as it is reasonable to expect that *N. gonorrhoeae* develops resistance to the current first-line drug during that time period (ceftriaxone started to be used as the first-line treatment for the US MSM in 2004 [14]). The total QALYs and costs were discounted to 2017 at 3% annually [15].

### Sensitivity analysis

We conducted one-way sensitivity analysis to investigate the impact of uncertainty in the utilities, durations and probabilities on our conclusions. The low and high parameter values from the associated uncertainty intervals were used.

## Results

The results of cost-effectiveness analysis are shown in Table 3. In the simulated cohort of 10,000 US MSM, performing the current GISP surveillance strategy results in $3.43M (95% uncertainty interval: $1.54M, $5.37M) and 32.5 (8.7, 66.9) QALYs over 25 years. Performing the eGISP strategy instead would results in $3.38M ($1.53M, $5.37M) and 31.9 (8.6, 66.9) QALYs.

**Table 3.** Results of cost-effectiveness analysis. The mean and 95% uncertainty intervals are reported. The simulations were run in the cohort of 10,000 US MSM over a 25-year period. The GISP strategy was compared with no surveillance, and the eGISP strategy was compared with the GISP strategy.

| Scenario | Total costs<br>(millions USD) | Total QALYs | Incremental costs (USD) | Incremental<br>QALYs gained | Outcome |
| --- | --- | --- | --- | --- | --- |
| No surveillance | 6.38 (2, 14.5) | 63.8 (12, 206.9) |  |  | Dominated |
| GISP surveillance strategy | 3.43 (1.54, 5.37) | 32.5 (8.7, 66.9) | -2.9M (-9.4M, -23,131) | 31.3 (0, 134.9) | Cost saving |
| eGISP surveillance strategy | 3.38 (1.53, 5.37) | 31.9 (8.6, 66.9) | -57,449 (-221,663, 100,914) | 0.59 (-0.79, 2.5) | Cost saving |

The current GISP surveillance strategy is cost saving compared to no surveillance as the costs are lower and the health gains are higher. However, it is dominated by eGISP surveillance strategy for the same reason.

Decomposition of the total discounted QALYs and costs over 25 years is shown in Figure 1.

**Figure 1.**
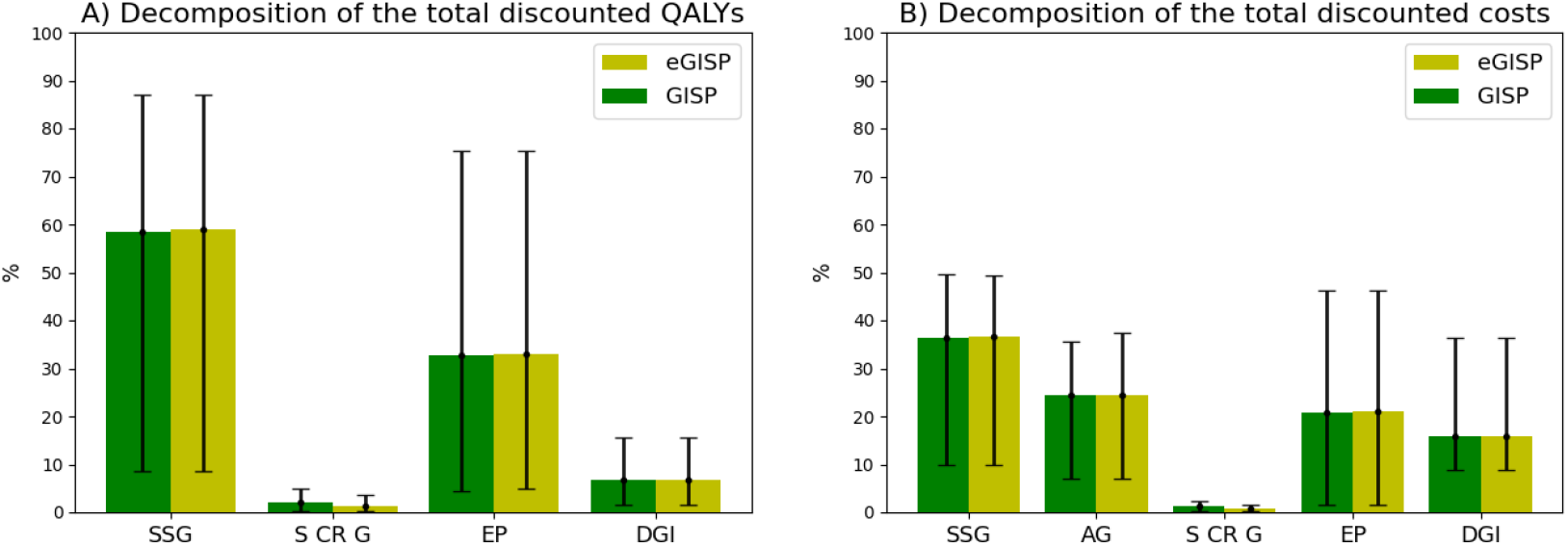
Decomposition of the total discounted QALYs and costs due to different types of gonococcal infection and its sequalae over 25 years under GISP and eGISP surveillance strategy. Means are identified as histograms, and 95% uncertainty intervals are identified as error bars. Abbreviations: SSG, symptomatic susceptible gonorrhea; AG, asymptomatic gonorrhea; S CR G, symptomatic ceftriaxone-resistant gonorrhea; EP, epididymitis; DGI, disseminated gonococcal infection.

Under GISP strategy, the composition of the total QALYs was 58.5% (8.6%, 87.1%) due to symptomatic susceptible gonorrhea, 2.2% (0.3%, 4.9%) due to symptomatic ceftriaxone-resistant gonorrhea, 32.8% (4.5%, 75.3%) due to epididymitis and 6.7% (1.6%, 15.7%) due to DGI. The contributors to the total costs were diagnosis and treatment of symptomatic susceptible gonorrhea (36.4% (10%, 49.5%)), diagnoses and treatment of asymptomatic gonorrhea (24.3% (7%, 35.5%)), diagnosis and treatment of symptomatic ceftriaxone-resistant gonorrhea (1.2% (0.3%, 2.4%)), treatment of epididymitis (20.9% (1.5%, 46.3%)) and treatment of DGI (15.9% (8.9%, 36.4%)).

Overall, the percentage of different types of gonococcal infection and the sequalae of the total QALYs and costs under eGISP strategy was very similar to the results obtained under GISP strategy. The contribution of symptomatic ceftriaxone-resistant gonorrhea into the disease burden under eGISP strategy was slightly lower than under GISP strategy (1.4% (0.17%, 3.6%) of the total QALYs and 0.8% (0.2%, 1.6%) of the total costs). Asymptomatic gonorrhea impacts directly in terms of costs but does not in terms of QALYs. The contribution of epididymitis into the disease burden is high overall. DGI impacts more in terms of costs than in terms of QALYs, accounting for over 15% of the total costs under both surveillance strategies. This complication is rare, but, unlikely epididymitis, has a nearly 30% probability of inpatient treatment resulting in high costs.

The results of one-way sensitivity analysis are presented in Figure 2. The exact values and the associated 95% uncertainty intervals are provided in Tables A-D in the supplement.

**Figure 2.**
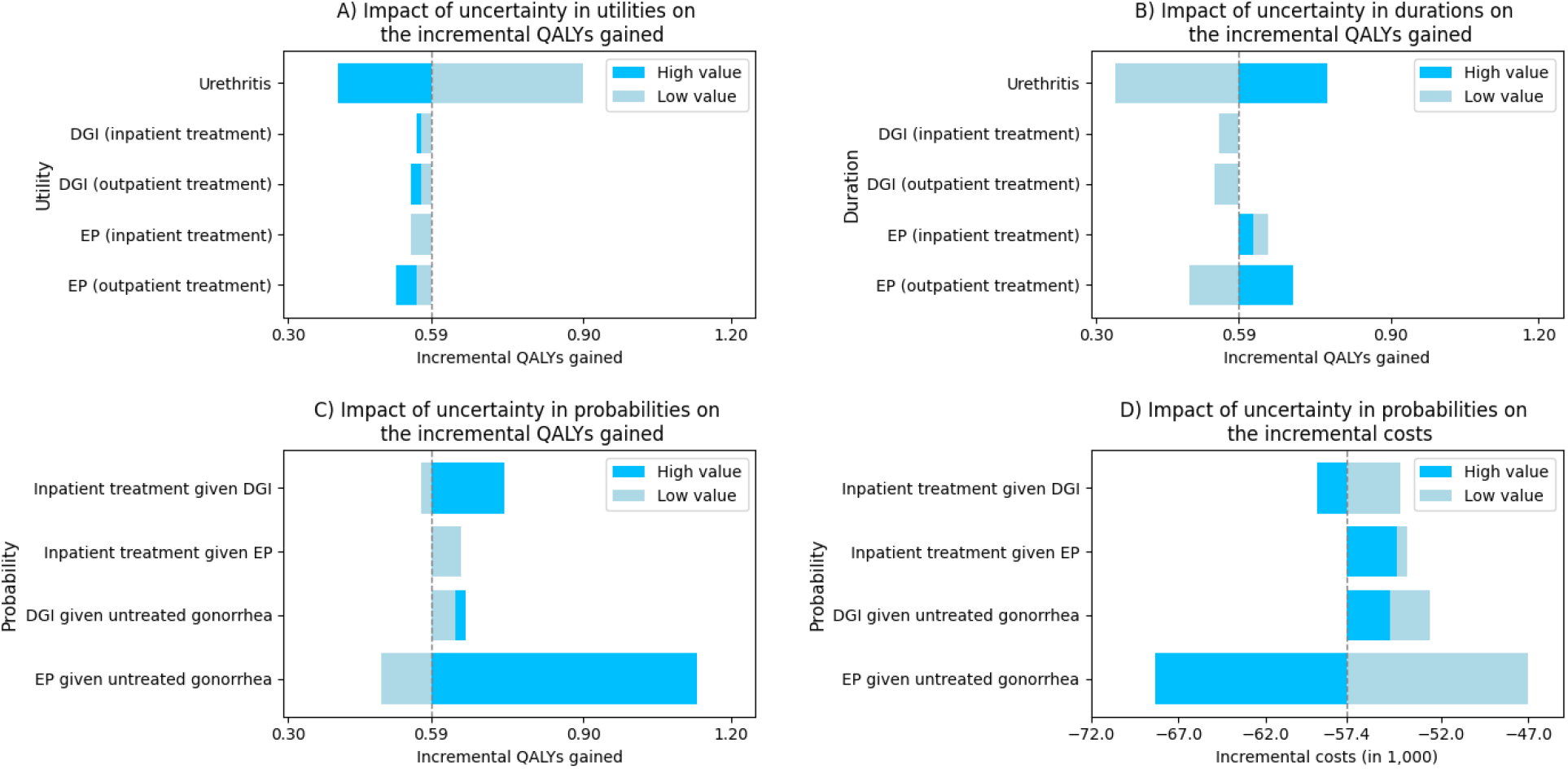
Results of one-way sensitivity analysis. In case the result obtained using a low value equals to the result obtained using a high value, only the low-value result is shown. Abbreviations: EP, epididymitis; DGI, disseminated gonococcal infection.

The eGISP surveillance strategy was cost saving compared to GISP strategy in all the one-way sensitivity analyses. The main determinants of the incremental QALYs gained were probability of epididymitis given untreated gonorrhea, utility of urethritis and duration of urethritis, while the incremental costs were the most sensitive to probability of ependymitis given untreated gonorrhea.

## Discussion

This study is continuation of our earlier work [8] where we determined that utilizing eGISP estimates for informing the gonorrhea treatment guidelines results in lower number of gonorrhea cases over a 25-year period without reduction in the lifespan of the current first-line antibiotic. In this paper, we estimated the total number of discounted QALYs and total discounted costs under both surveillance strategies over the same period of time. Our results indicate that performing the surveillance of AMR gonorrhea under the current GISP surveillance strategy is cost saving compared to no surveillance. However, switching from the current strategy to eGISP strategy is expected to further reduce the costs and increase the health gains.

Overall, there was no significant difference in the contribution of different types of gonococcal infection and the sequalae into the disease burden under both surveillance strategies. However, as expected, the contribution of symptomatic ceftriaxone-resistant gonorrhea was slightly higher under GISP strategy than under eGISP strategy. Symptomatic susceptible gonorrhea is the main source of the total QALYs and costs contributing for around 60% and 35% of them, respectively.

The results that we obtained for contribution of the sequalae into the gonorrhea burden are significantly higher than the ones reported in the earlier study [11]. This is probably because in [11] it was assumed that only untreated urethral infections can lead to development of the sequalae. In our modelling study, the sequalae can occur as the result of untreated gonorrhea at any affected site (urethra, rectum and pharynx) which results in more realistic estimates. The significant contribution of the sequalae into the disease burden once again highlights the importance of routine screening for sexually transmitted infections which is currently believed to be significantly below the recommended level for the US MSM [16].

This analysis should be viewed in the context of some limitations. We did not account for the increased ability to acquire and transmit the HIV infection in case of gonococcal infection. Also, a full adherence to the gonorrhea treatment guidelines was assumed. In reality, according to the recent study [17], around 20% of the male patients have not received the recommended first-line antibiotic therapy. Probability of epididymitis in case of untreated gonococcal infection obtained from [11] was based on pooled estimates from longitudinal studies on sequelae among men with chlamydia as no data for gonorrhea was available. If such data become available in the future, it will improve the accuracy of this analysis.

The goal of establishment of eGISP was to study whether there is a difference in antibiotic susceptibility pattern between men and women and to test the assumption whether rectum and pharynx can serve as a niche for the growth of antibacterial resistance [19]. Its goal has never been to inform the gonorrhea treatment guidelines as GISP has been traditionally used for this purpose. Our findings provide a new perspective. We demonstrated that switching from the current surveillance strategy to eGISP strategy is cost saving and should be considered in order to improve the population health and reduce the financial burden of gonorrhea.

## Data availability statement

All relevant data are within the manuscript and its supplement.

## References

1. Alirol, E., et al., Multidrug-resistant gonorrhea: A research and development roadmap to discover new medicines. PLoS medicine, 2017. 14(7): p. e1002366.

2. Workowski, K., Chlamydia and gonorrhea. Annals of internal medicine, 2013. 158(3): p. ITC2–1.

3. Maurakis, S.A. and C.N. Cornelissen, Recent progress towards a gonococcal vaccine. Frontiers in cellular and infection microbiology, 2022. 12: p. 881392.

4. Gaydos, C.A. and J.H. Melendez, Point-by-point progress: gonorrhea point of care tests. Expert review of molecular diagnostics, 2020. 20(8): p. 803–813.

5. Bolan, G.A., P.F. Sparling, and J.N. Wasserheit, The emerging threat of untreatable gonococcal infection. The New England journal of medicine, 2012. 366(6): p. 485.

6. Organization, W.H., Global action plan to control the spread and impact of antimicrobial resistance in Neisseria gonorrhoeae. 2012: World Health Organization.

7. Pham, C., et al., Gonococcal isolate surveillance project (GISP) and enhanced GISP (eGISP). 2020.

8. Prakhova, S., Evaluating Performance of the US Surveillance Systems for Monitoring Antimicrobial-Resistant Gonorrhea: An Agent-Based Modelling Study. 2024.

9. Husereau, D., et al., Consolidated Health Economic Evaluation Reporting Standards 2022 (CHEERS 2022) statement: updated reporting guidance for health economic evaluations. MDM Policy & Practice, 2022. 7(1): p. 23814683211061097.

10. Li, Y., et al., The estimated lifetime quality-adjusted life-years lost due to chlamydia, gonorrhea, and trichomoniasis in the United States in 2018. The Journal of Infectious Diseases, 2023. 227(8): p. 1007–1018.

11. Li, Y., et al., Estimated costs and quality-adjusted life-years lost due to N. gonorrhoeae infections acquired in 2015 in the United States: a modelling study of overall burden and disparities by age, race/ethnicity, and other factors. The Lancet Regional Health–Americas, 2022. 16.

12. Statistics, U.S.B.o.L. Consumer Price Index (CPI) databases. 2024; Available from: https://www.bls.gov/cpi/data.htm.

13. Ertapenem Prices, Coupons and Patient Assistance Programs. Available from: https://www.drugs.com/price-guide/ertapenem.

14. Del Rio, C., et al., Update to CDC’s sexually transmitted diseases treatment guidelines, 2006: fluoroquinolones no longer recommended for treatment of gonococcal infections. JAMA: Journal of the American Medical Association, 2007. 297(22).

15. Lopez, A.D. and C.C. Murray, The global burden of disease, 1990–2020. Nature medicine, 1998. 4(11): p. 1241–1243.

16. Earnest, R., et al., Population-level benefits of extragenital gonorrhea screening among men who have sex with men: an exploratory modeling analysis. Sexually transmitted diseases, 2020. 47(7): p. 484–490.

17. Sittig, K.R., S.M. Collin, and R. Rosa, Factors associated with non-guideline-adherent treatment for gonorrhea and chlamydia among outpatient prescriptions in the Unites States. International journal of STD & AIDS, 2022. 33(7): p. 694–700.

18. Control, C.f.D., CDC No Longer Recommends Oral Drug for Gonorrhea Treatment. Press Release, Aug, 2012. 9: p. 1.

19. St Cyr, S., et al., Gonococcal Isolate Surveillance Project (GISP) and Enhanced GISP (eGISP) Protocol. 2021.

