## Supplement for "Cost-effectiveness of the Strategies for Surveillance of Antimicrobial-resistant Gonorrhea in the US: a Modelling Study"

### Supplementary material

#### 1. Calculation of costs

The cost of diagnosis and treatment of a symptomatic susceptible case ($Cost_{ssg}$) includes the cost of testing for gonorrhea ($c_{gt}$), cost of the current first-line antibiotic for gonorrhea ($c_{at1}$) and cost of treatment of urethritis ($c_U$) which includes the cost of a short clinic visit (in consistence with the previous studies [1] [2]).

$Cost_{ssg}$=$c_{gt} + c_{at1} + c_U$.

In 2017-2020, a combination of ceftriaxone 250g and azithromycin 1g was the first-line therapy [3]. In 2020, it was replaced by ceftriaxone 500g [3]. We assumed that its cost as well as the cost of an antibiotic that replaces ceftriaxone after the switch equals to the double cost of ceftriaxone 250g.

The cost of diagnosis and treatment of a detected asymptomatic case ($Cost_{ag}$) consists of almost the same components as the cost of diagnosis and treatment of a symptomatic susceptible case, expect for the cost of a short clinic visit ($c_{sv}$) used instead of the cost of treatment of urethritis. For the US men who have sex with men (MSM), the routine screening is recommended by the Centers for Disease Control and Prevention (CDC) guidelines [4] at the sites for which a patient reported the exposure. We assumed that the majority of the MSM were exposed at all three anatomical sites since their last screening. However, it is widely believed that the CDC recommendations are not always followed by the healthcare professionals [5]. Therefore, we assigned the number of sites for which a screening was performed by *n*=uniform (1, 3).

$Cost_{ag}$=$nc_{gt} + c_{at1} + c_{sv}$.

The cost of diagnosis and treatment of symptomatic ceftriaxone-resistant case ($Cost_{scrg}$) includes the cost of diagnosis and treatment of a symptomatic susceptible case ($Cost_{ssg}$), cost of re-treatment with the second-line drug ($c_{at2}$) and cost of treatment of urethritis ($c_U$).

$Cost_{scrg}$=$Cost_{ssg} + c_{at2} + c_U$.

The second-line antibiotic is ertapenem 1g. The cost of antimicrobial susceptibility testing (AST) upon patient return to a healthcare provider with suspected treatment failure was not included as currently the Maryland and Washington State Public Health Labs offer nationwide AST at no cost through the CDC's Antimicrobial Resistance Laboratory Network [7].

The cost of treatment of a sequala (epididymitis or disseminated gonococcal infection (DGI)) per case of untreated gonococcal infections was calculated as:

$Cost_{seq_i} = p_i(c_{ni}p_{ni} + c_{oi}(1 - p_{ni}))$,

$i = 1,2$,

where $p_i$ is the probability of the sequala $i$ given untreated gonococcal infection; $p_{ni}$ is the probability of inpatient treatment given sequala $i$; $c_{ni}$ and $c_{oi}$ is the cost of inpatient and outpatient treatment of sequala $i$, respectively.

#### 2. Results of sensitivity analysis

| Utilities | Incremental QALYs gained | |
|---|---|---|
| | Low value | High value |
| Urethritis | 0.9 (-0.8, 4.3) | 0.4 (-0.5, 2.9) |
| DGI (inpatient treatment) | 0.57 (-0.68, 2.22) | 0.56 (-0.65, 2.24) |
| DGI (outpatient treatment) | 0.57 (-0.68, 2.21) | 0.55 (-0.64, 2.25) |
| Epididymitis (inpatient treatment) | 0.55 (-0.56, 2.26) | 0.55 (-0.56, 2.26) |
| Epididymitis (outpatient treatment) | 0.56 (-0.79, 2.25) | 0.52 (-0.52, 2.28) |

**Table A. Impact of uncertainty in utilities on the incremental QALYs gained.** The mean and 95% uncertainty intervals are reported. The simulations were performed using the low and high parameter values from the corresponding uncertainty intervals in Table 1 in the main text. Abbreviation: DGI, disseminated gonococcal infection.

| Durations | Incremental QALYs gained | |
|---|---|---|
| | Low value | High value |
| Urethritis | 0.34 (-0.52, 1.54) | 0.77 (-1.14, 3.33) |
| DGI (inpatient treatment) | 0.55 (-0.55, 2.28) | 0.55 (-0.56, 2.25) |
| DGI (outpatient treatment) | 0.54 (-0.55, 2.29) | 0.56 (-0.57, 2.25) |
| Epididymitis (inpatient treatment) | 0.65 (-0.49, 3) | 0.62 (-0.5, 3) |
| Epididymitis (outpatient treatment) | 0.49 (-0.44, 2.29) | 0.7 (-0.63, 3.56) |

**Table B. Impact of uncertainty in durations on the incremental QALYs gained.** The mean and 95% uncertainty intervals are reported. The simulations were performed using the low and high parameter values from the corresponding uncertainty intervals in Table 1 in the main text. Abbreviation: DGI, disseminated gonococcal infection.

| Probabilities | Incremental QALYs gained | |
|---|---|---|
| | Low value | High value |
| Probability of inpatient treatment given DGI | 0.57 (-1.8, 3.18) | 0.74 (-0.78, 3) |
| Probability of inpatient treatment given epididymitis | 0.65 (-0.5, 3) | 0.65 (-0.5, 3) |
| Probability of DGI given untreated gonorrhea | 0.64 (-0.49, 2.99) | 0.66 (-0.53, 3) |
| Probability of epididymitis given untreated gonorrhea | 0.49 (-0.49, 2.28) | 1.13 (-1.64, 10.19) |

**Table C. Impact of uncertainty in probabilities on the incremental QALYs gained.** The mean and 95% uncertainty intervals are reported. The simulations were performed using the low and high parameter values from the corresponding uncertainty intervals in Table 1 and 2 in the main text. Abbreviation: DGI, disseminated gonococcal infection.

| Probabilities | Incremental costs (USD) | |
|---|---|---|
| | Low value | High value |
| Probability of inpatient treatment given DGI | -54,312 (-197,744, 100,341) | -59,132 (-252,139, 112,121) |
| Probability of inpatient treatment given epididymitis | -53,908 (-206,641, 108,027) | -54,515 (-210,073, 109,392) |
| Probability of DGI given untreated gonorrhea | -52,592 (-199,505, 104,154) | -54,890 (-223,790, 115,594) |
| Probability of epididymitis given untreated gonorrhea | -47,066 (-176,215, 93,500) | -68,341 (-456,958, 146,221) |

**Table D. Impact of uncertainty in probabilities on the incremental costs.** The mean and 95% uncertainty intervals are reported. The simulations were performed using the low and high parameter values from the corresponding uncertainty intervals in Table 1 and 2 in the main text. Abbreviation: DGI, disseminated gonococcal infection.

#### References


1. Mehta, S.D., et al., *Cost-effectiveness of five strategies for gonorrhea and chlamydia control among female and male emergency department patients.* Sexually transmitted diseases, 2002. **29**(2): p. 83-91.
2. Li, Y., et al., *Estimated costs and quality-adjusted life-years lost due to N. gonorrhoeae infections acquired in 2015 in the United States: a modelling study of overall burden and disparities by age, race/ethnicity, and other factors.* The Lancet Regional Health–Americas, 2022. **16**.
3. Cyr, S.S., *Update to CDC's treatment guidelines for gonococcal infection, 2020.* MMWR. Morbidity and mortality weekly report, 2020. **69**.
4. Workowski, K.A., *Sexually transmitted infections treatment guidelines, 2021.* MMWR. Recommendations and Reports, 2021. **70**.
5. Earnest, R., et al., *Population-level benefits of extragenital gonorrhea screening among men who have sex with men: an exploratory modeling analysis.* Sexually transmitted diseases, 2020. **47**(7): p. 484-490.
6. Li, X., et al., *In vitro activity of ertapenem against Neisseria gonorrhoeae clinical isolates with decreased susceptibility or resistance to extended-spectrum Cephalosporins in Nanjing, China (2013 to 2019).* Antimicrobial Agents and Chemotherapy, 2022. **66**(5): p. e00109-22.
7. Prevention, C.f.D.C.a., *Gonorrhea Laboratory Information.* 2021.
